# ANAEMIA IN CHILDREN AGED 7 MONTHS TO 12 YEARS AND ASSOCIATED FACTORS: A CROSS-SECTIONAL STUDY OF CHILDREN ADMITTED AT THE EFFIDUASE DISTRICT HOSPITAL AND AHMADIYYA MISSION HOSPITAL, GHANA

**DOI:** 10.1101/2023.04.25.23289117

**Authors:** Antwi Joseph Barimah, Mohammed Mohammed Ibrahim, Solomon Saka Allotey, Bernard Opoku Amoah, Larry Agyemang, James Dumba, Yaw Boakye Nketiah, Rebecca Dorcas Commey, Semefa Alorvi, Deborah Ampofo

**Author notes:** Corresponding Author Antwi Joseph Barimah. **Declaration of interests** The authors declare no competing interests. **Author contributions** All authors contributed significantly to the critical revision and approved the final version before the onward submission. **Funding** This study received no funding.

## Abstract

A number of factors have been identified as influencing the prevalence of anaemia in children. In the Sekyere East district, an increasing trend of children with severe anaemia leading to haemotransfusion has been observed. Over a three-month period (June to August 2022) approximately one hundred (100) children were haemotransfused due to severe anaemia in the Effiduase district hospital. This quantitative oriented study adopted the descriptive cross-sectional study design. Specifically, the study sought to explore the prevalence of childhood anaemia and its associated factors in the district. Purposive sampling technique which is a non-probability (non-randomized) was used to select children and their guardians to partake in the study. SPSS statistical software version 25 was used to analyze quantitative data of this study and data were presented in frequencies using tables. This study has accentuated high prevalence of anaemia in the Sekyere East district looking at the various findings brought to light by the Haemoglobin (Hb) blood analysis, Mean Corpuscular Volume (MCV) blood tests and the Mean Corpuscular Haemoglobin (MCH) test results. The findings from the study have also shown that malaria in children is significantly associated with the onset of childhood anaemia. The study additionally revealed moderate nutritional intake of food products very rich in iron, vitamin B12 and folate nutrients necessary to curb anaemia in children. In conclusion, study findings therefore underscore the need for multi-faceted approaches that address both malaria control and nutrition in order to reduce anaemia among the children in the Sekyere East district.

## 1.0 INTRODUCTION

Micronutrient deficiencies impose substantial health, economic, and social burdens worldwide. Iron deficiency (ID) is the most prevalent haematologic disorder during childhood, globally. One major consequence of ID is anaemia, which affects a large proportion of the world’s population. ^1^ Anaemia has a range of adverse consequences including; poor cognitive performance, poor growth of infants, preschool and school-aged children; impairment of physical capacity and work performance of adolescents and adults; reduction in immune competence; and increased morbidity from infections in all age groups.^8^ Severe anaemia is a common condition causing significant morbidity and mortality in children in Africa. Severe anaemia carries a high “hidden” morbidity and mortality occurring in the months after initial diagnosis and treatment.^2^

Anaemia in children is a public health issue in Ghana. It has been reported that 76% of Ghanaian children below five years of age, 73% of children aged 2-10 years, and 63% of school children aged 5-12 years suffer from anaemia.^3^ Anaemia has multiple causes and associated risk factors which often work in tandem. They include various nutritional deficiencies, infections and infestations, as well as genetic defects including glucose-6-phosphate dehydrogenase (G6PD) deficiency, and haemoglobinopathies. Socio-economic and cultural factors like poverty, illiteracy, ignorance, superstition and cultural beliefs and practices also influence the incidence of anaemia, most of them indirectly through poor nutrition.^4^ Studies in Kenya, India and Bangladesh have shown multiple coexisting causes of anaemia in individuals.^9^ These causes include iron deficiency, vitamin A deficiency, malaria parasitaemia and other parasitic infestations. Traditionally, haemoglobin level has been used to estimate iron deficiency and iron deficiency anaemia even though haemoglobin estimates are neither specific nor sensitive as a screening test for iron.^5^ Across the spectrum of anaemia, iron deficiency anaemia (IDA) is the most prevalent nutritional disorder worldwide accounting for about 75%-80% of the total burden of anaemia. IDA is partly induced by plant-based diets containing low bioavailable non-haeme iron. Blood losses within the gastrointestinal tract due to intestinal parasites or inflammatory bowel disease may also contribute to IDA.^6^

In the Sekyere East district, an increasing trend of children with severe anaemia leading to haemotransfusion has been observed. Over a three months period (June to August 2022) approximately one hundred (100) children were haemotransfused due to severe anaemia in the Effiduase district hospital. This is a cause for concern given the acute and long-term negative impact associated with anaemia especially amongst children.^7^

However, the major contributing factors of anaemia among Ghanaian children aged 7 months to 12 years are largely understudied. Existing studies on anaemia focus primarily on preschool children (< 5 years) and little is known about anaemia in school-aged children in rural Ghana. The objective of the current study was to assess the situation of anaemia and the possible risk factors in the Sekyere east district of Ghana.

## 2.0 MATERIALS AND METHODS

### 2.1 Study Area

The study was carried out in the Sekyere East District of the Ashanti Region of Ghana. The District created in 1988 has Effiduase as its capital, it is one of the 30 districts in the Ashanti Region of Ghana. The population of the district according to the 2010 population and housing census stands at 62,172 with 29,511 males 32,661 females and 23,320 been between the age of 0-14years. There are eight (8) health facilities in the District made up of two (2) hospitals, five (5) clinics and one (1) CHPS Compound. Three of these facilities are public facilities (the Effiduase District Hospital, Okaikrom Health Centre and the Aherewa CHPS Compound), while the remaining five are owned by the Ahmadiyya Mission, the Methodist Mission, the Catholic Church and two (2) privately owned facilities (1 hospital & 1clinic) according to the Ghana Statistical Service, 2014.

### 2.2 Research Design and Study Population

The study utilized a descriptive cross-sectional approach in exploring the prevalence of anaemia among children aged 7months -12years. The study subjects were children on admission at the children’s ward of the two major hospitals in the district: Effiduase District Hospital in Effiduase and Ahmadiyya Missions Hospital at Asokore.

### 2.3 Sample Size and Sampling Technique

The study utilized a sample size of 210 children between 7months -12 years admitted between August and November 2022. Hence, the sample size of 210 was arrived at as a result of the children admitted between the period of August and November 2022. Purposive sampling was used to include children who met the inclusion criteria. Purposive sampling was utilized because of the peculiar nature of the study participants, hence, children who met the criteria being sought by authors were purposively selected.

### 2.4 Data Collection Instrument

Questionnaire was used to collect data for this current study, the instrument was structured based on the objectives of the study outlined earlier. The content of the instrument included a child’s demographic characteristics, guardian’s information, general medical history, laboratory information, haemotransfusion background and questions pertaining to risk factors. The instrument contained both open ended and close ended questions.

### 2.5 Data Collection Procedure

#### 2.5.1 Clinical Data

Data was collected with the help of a questionnaire which was administered to care givers and filled by designated staff members at the children’s ward within 24hours of admission. The data was collected from the 210 children between 7months -12 years admitted between August and November 2022 at the Effiduase District Hospital and Ahamadiyya Mission Hospital. Since clinical data was captured from the medical records book (folder) of children which contained their sociodemographic characteristics, authors could assess information that could identify individual participants during data collection. However, none of the data was abused as authors ensured optimum privacy and confidentiality of study participants as enshrined in the ethical principles.

#### 2.5.2 Laboratory Data

For each of the study participants, samples were taken for Full Blood Count investigation on admission before any intervention to correct anaemia was initiated. The results of the FBC was recorded onto the questionnaire. Such data was collected from the 210 children between 7months -12 years admitted between August and November 2022 at the Effiduase District Hospital and Ahamadiyya Mission Hospital. Confidentiality of such data was highly ensured as the identity was the case during the capturing of the clinical data by authors.

### 2.6 Data Analysis

Statistical Package for the Social Sciences (SPSS version 25) is the data analysis tool that was used to analyze data of this study. Questionnaires after being collected was thoroughly checked through to ensure all questions were accurately answered and errors were corrected to ensure completeness before entering it into the SPSS for rigorous data analysis.

### 2.7 Ethical Considerations

Since the study involved the use of human subjects, ethical approval was sought from Ghana Health Service Ethical Review Committee who rigorously scrutinized the protocols before finally granting an ethical backing to the study. Permission was further sought from the administrations of the Effiduase District Hospital and The Ahmadiyya Mission Hospital before the commencement of the study. Informed consent was also sought from the care givers of children between 7months and 12 years on admission at the selected facilities. They were assured of optimum privacy and confidentiality of information given during the conduct of the study.

## 3.0 RESULTS

### 3.1 Presentation of results

From the table 1 above, a little above half of the children were males (56.2%) and 43.8% were females. Children between 2years-5years dominated with 40.5%, 7months-2years occupied the next higher frequency with 40% and 19.5% representing the least category were between the ages of 5years-12years. On the sub-district of children, Asokore dominated with 38.6%, followed by Effiduase with 29%, Others who were mostly from Kumawu occupied the next frequency with 24.3%, followed by Mponua with 5.2% and 2.9% representing the least sub-district was Nyanfa. On the number of siblings, children with no sibling dominated with 22.4%, followed by children with 1 and 2 siblings who occupied the next level with 21%, this was followed by 18.6% who had 3 siblings, 9% of the children had 5 or more siblings and the least category were children with 4 siblings 8.1%. On birth placement, 30.5% of the children were first born, 22.4% of the children were second born, 21% were third born, 15.2% were fifth or more born and the least were 4 born with 11%.

**Table 1:** Background Attributes of Children (N=210)

| Attributes | Frequency | Percentage % |
| --- | --- | --- |
| **Gender** |  |  |
| Male | 118 | 56.2 |
| Female | 92 | 43.8 |
| **Age** |  |  |
| 7months-2years | 84 | 40.0 |
| 2years-5years | 85 | 40.5 |
| 5years-12years | 41 | 19.5 |
| **Sub-district** |  |  |
| Effiduase | 61 | 29.0 |
| Asokore | 81 | 38.6 |
| Nyanfa | 6 | 2.9 |
| Mponua | 11 | 5.2 |
| Others | 51 | 24.3 |
| **Number of sibling** |  |  |
| 0 | 47 | 22.4 |
| 1 | 44 | 21.0 |
| 2 | 44 | 21.0 |
| 3 | 39 | 18.6 |
| 4 | 17 | 8.1 |
| >5 | 19 | 9.0 |
| **Birth placement** |  |  |
| 1<sup>st</sup> | 64 | 30.5 |
| 2<sup>nd</sup> | 47 | 22.4 |
| 3<sup>rd</sup> | 44 | 21.0 |
| 4<sup>th</sup> | 23 | 11.0 |
| 5th or more | 32 | 15.2 |

***Source: Field Survey, 2022***

From table 2 above, a greater majority of the guardians were mothers of children with 81.4%, this was followed by other guardians with 12.4%, fathers of children occupied the next level with 4.8% and the least guardians were siblings of children with 1.4%. On the educational level of guardians, a little above half of guardians were basic school leavers, 21% of guardians had no formal education, 11.9% were secondary school leavers, 7.1% of guardians had tertiary education and a least number of guardians 1.4% had post-secondary school education. On religion of guardians, Christians dominated with 84.8%, Muslims were 14.8% and 0.5% were Traditionalists. On employment of guardians, a good majority of them (78.1%) were employed while 21.9% were unemployed. A huge majority of the guardians (92.4%) indicated that their household heads were employed, 4.8% indicated their household heads were unemployed and 2.9% gave no response to the aforementioned question. On the marital status of guardians, more than half of the guardians (63.3%) were married, 12.9% indicated they were single and co-habiting, 7.1% were divorced while 3.8% were widowed.

**Table 2:** Guardians information (N=210)

| Attributes | Frequency | Percentage % |
| --- | --- | --- |
| **Guardians relationship** |  |  |
| Mother | 171 | 81.4 |
| Father | 10 | 4.8 |
| Sibling | 3 | 1.4 |
| Other | 26 | 12.4 |
| **Educational level** |  |  |
| None | 44 | 21.0 |
| Basic | 123 | 58.6 |
| Secondary | 25 | 11.9 |
| Post-secondary | 3 | 1.4 |
| Tertiary | 15 | 7.1 |
| **Religion** |  |  |
| Christian | 178 | 84.8 |
| Muslim | 31 | 14.8 |
| Traditional | 1 | .5 |
| **Employment of guardian** |  |  |
| Employed | 164 | 78.1 |
| Unemployed | 46 | 21.9 |
| **Employment of household head** |  |  |
| Employed | 194 | 92.4 |
| Unemployed | 10 | 4.8 |
| No response | 6 | 2.9 |
| **Marital status** |  |  |
| Married | 133 | 63.3 |
| Single | 27 | 12.9 |
| Divorced | 15 | 7.1 |
| Widowed | 8 | 3.8 |
| Co-habiting | 27 | 12.9 |

**Source: Field Survey, 2022**

Table 3 vividly highlights the medical history of children as at the time of the survey, the study observed that a good majority of the children (93.3%) had a normal weight of >2.5kg while 6.7% had low weight <2.5kg. On the immunization status of children, a huge majority of the children (91.9%) had up to date immunization while a few 8.1% had defaulted in one way or the other. Amongst the defaulters, majority defaulted at 18months, followed by (1 year two months, 36 months, 24 months and 12 months) occupied the next level with the same frequency. On the growth monitoring chart, the study observed that a good majority of the children (94.3%) had normal growth, 4.8% had a plateau growth (unstable) while 1% had a declining growth.

**Table 3:** Medical history (N=210)

| Attributes | Frequency | Percentage % |
| --- | --- | --- |
| **Birth weight** |  |  |
| Normal weight (>2.5kg) | 196 | 93.3 |
| Low weight (<2.5kg) | 14 | 6.7 |
| **Immunization** |  |  |
| Up to date | 193 | 91.9 |
| Defaulted | 17 | 8.1 |
| **Defaulted/compliance** |  |  |
| 12 MONTHS | 1 | .5 |
| 18 MONTHS | 11 | 5.2 |
| 24 MONTHS | 1 | .5 |
| 36 MONTHS | 1 | .5 |
| ONE YEAR | 2 | 1.0 |
| ONE YEAR,TWO MONTHS | 1 | .5 |
| Not applicable | 193 | 91.9 |
| **Growth chart** |  |  |
| Normal | 198 | 94.3 |
| Plateau | 10 | 4.8 |
| Decline | 2 | 1.0 |
| **Breast fed** |  |  |
| Yes | 208 | 99.0 |
| No | 2 | 1.0 |
| **Type of breast feeding** |  |  |
| Exclusively | 44 | 21.0 |
| Partly | 164 | 78.1 |
| **Known chronic disease** |  |  |
| SCD | 7 | 3.3 |
| None | 203 | 96.7 |
| **Number of admission** |  |  |
| FIVE TIMES | 8 | 3.8 |
| FOUR TIMES | 7 | 3.3 |
| ONCE | 132 | 62.9 |
| THRICE | 18 | 8.6 |
| TWICE | 45 | 21.4 |
| **Dewormed over past year** |  |  |
| Yes | 87 | 41.4 |
| No | 123 | 58.6 |

***Source: Field Survey, 2022***

**Table 4:**
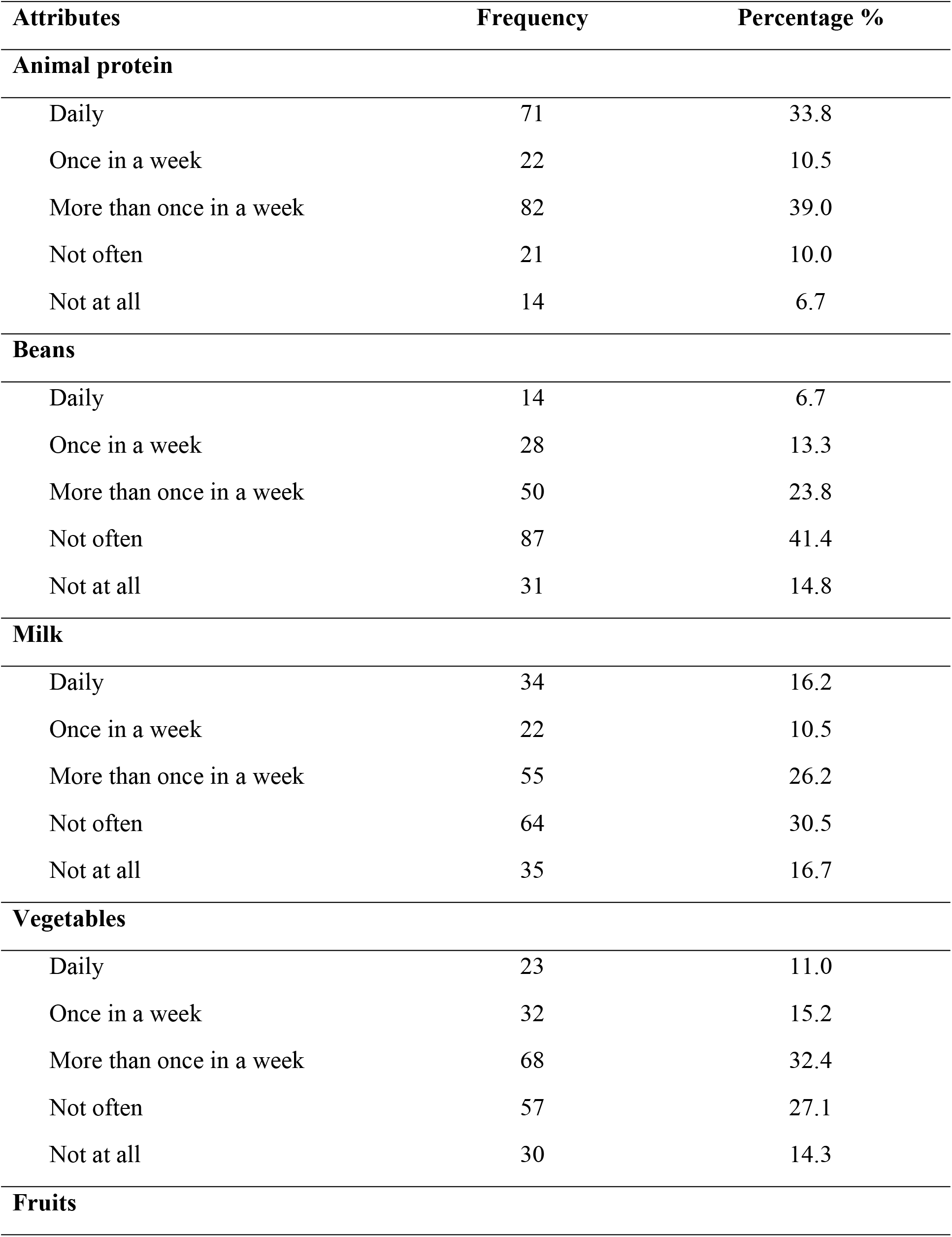
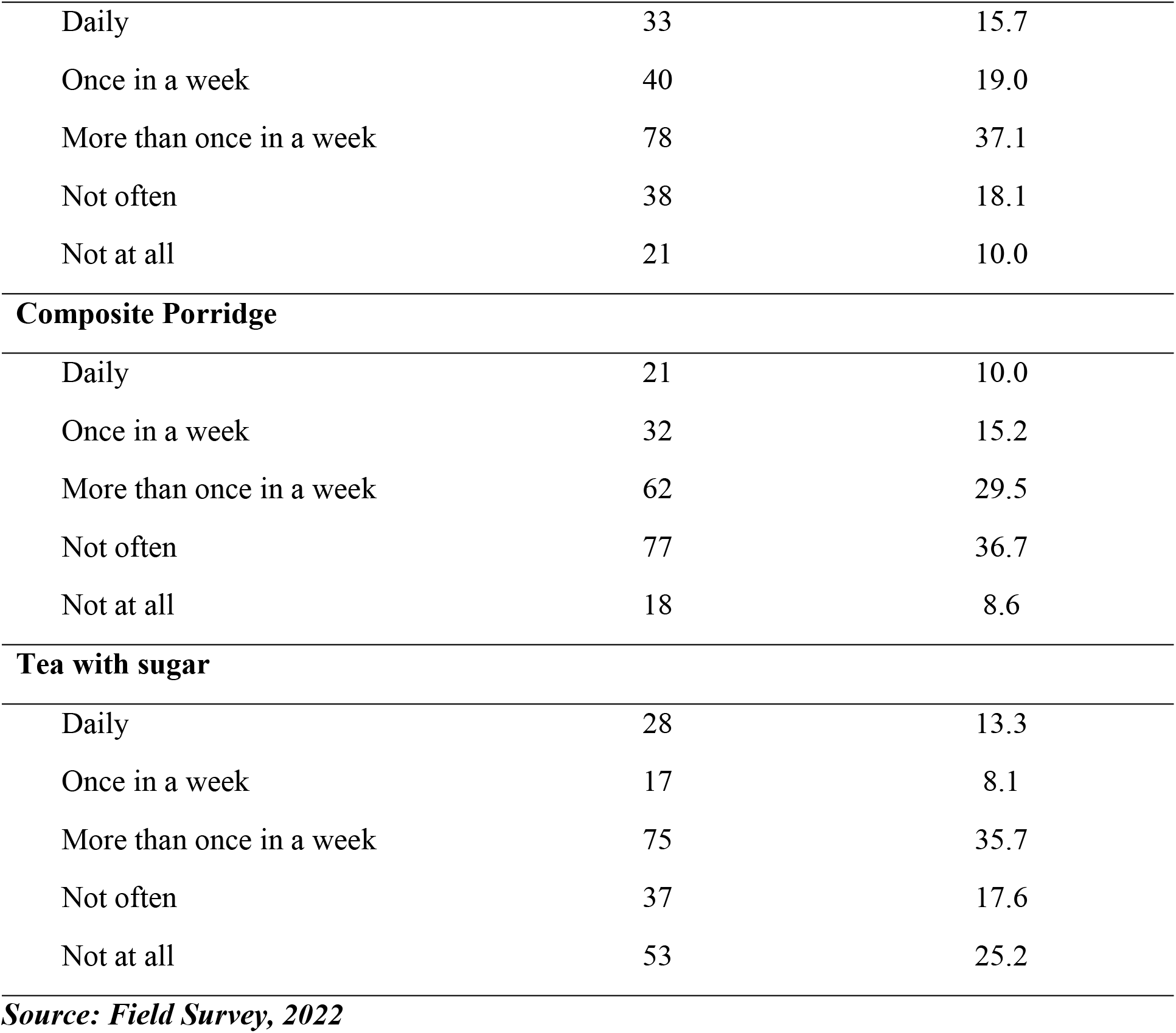
Risk factors (N=210)

**Table 5:** Principal Diagnosis (N=210)

| **Attributes** | **Frequency** | **Percentage %** |
| --- | --- | --- |
| Severe Anaemia | 15 | 7.1 |
| Mild Anaemia | 3 | 1.4 |
| Moderate Anaemia | 6 | 2.9 |
| Malaria | 42 | 20 |
| Severe Malaria | 63 | 30 |
| Parotitis | 1 | 0.5 |
| Febrile Illness | 6 | 2.9 |
| UTI | 2 | 1.0 |
| Mild anaemia and Sepsis | 1 | 0.5 |
| Complicated Malaria | 2 | 1.0 |
| Burns | 3 | 1.4 |
| Severe anaemia and Severe malaria | 1 | 0.5 |
| Sepsis | 5 | 2.4 |
| Febrile convulsion and Severe malaria | 5 | 2.4 |
| Gastroenteritis | 7 | 3.3 |
| Bronchitis | 3 | 1.4 |
| RTI | 2 | 1.0 |
| UTI and Malaria | 1 | 0.5 |
| Bronchopneumonia | 5 | 2.4 |
| Pneumonia | 2 | 1.0 |
| Moderate Anemia and Sepsis | 1 | 0.5 |
| Gastroenteritis and Malaria | 4 | 1.9 |
| Gastroenteritis and Complicated malaria | 1 | 0.5 |
| Sepsis and Malaria | 2 | 1.0 |
| Febrile Convulsion | 3 | 1.4 |
| Ineffective gastroenteritis | 1 | 0.5 |
| A.R.I | 1 | 0.5 |
| Left inguinal hernia | 2 | 1.0 |
| Moderate anaemia and Malaria | 2 | 1.0 |
| Allergic rhinitis | 1 | 0.5 |
| Severe malaria and Enteritis | 2 | 1.0 |
| Moderate anaemia and Severe malaria | 2 | 1.0 |
| Mild anaemia and Severe malaria | 3 | 1.4 |
| Nephrotic syndrome | 1 | 0.5 |
| Severe anaemia in SCD | 5 | 2.4 |
| Malaria in SCD | 1 | 0.5 |
| Sepsis in SCD | 1 | 0.5 |

***Source: Source: Field Survey, 2022***

**Table 6:**
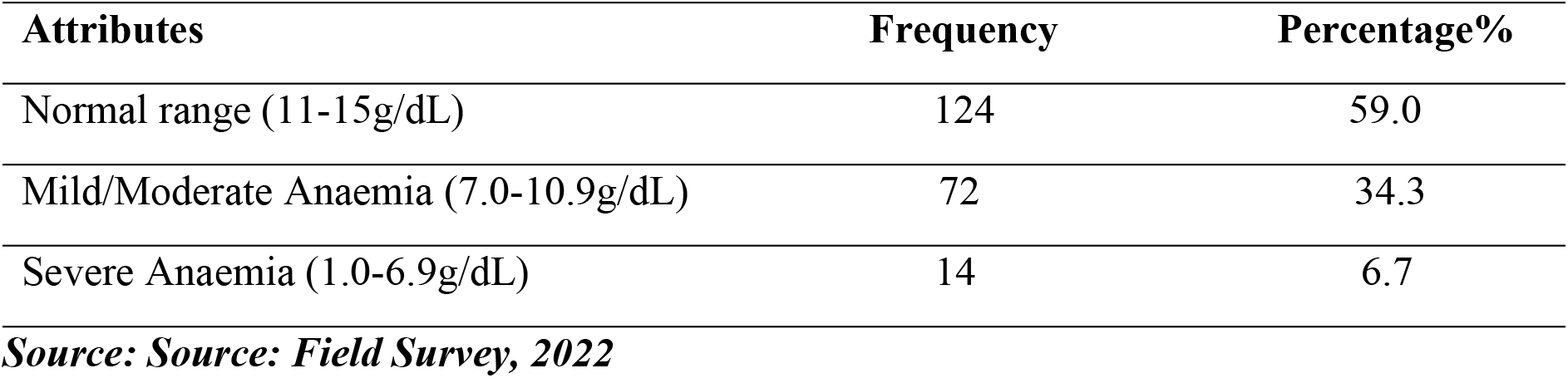
Laboratory investigation (Hb) (N=210)

| Attributes | Frequency | Percentage% |
| --- | --- | --- |
| Normal range (11-15g/dL) | 124 | 59.0 |
| Mild/Moderate Anaemia (7.0-10.9g/dL) | 72 | 34.3 |
| Severe Anaemia (1.0-6.9g/dL) | 14 | 6.7 |

**Source: Source: Field Survey, 2022**

**Table 7:** Laboratory investigation (MCV) (N=210)

| Attributes | Frequency | Percentage% |
| --- | --- | --- |
| Normal range (80-95FL) | 33 | 15.7 |
| Iron deficiency Anaemia (<80FL) | 176 | 83.8 |
| Vitamin B12 or Folate deficiency Anaemia (>95FL) | 1 | 0.5 |

**Source: Source: Field Survey, 2022**

**Table 8:** Laboratory investigation (MCH) (N=210)

| Attributes | Frequency | Percentage% |
| --- | --- | --- |
| Normal range (27-34PG) | 28 | 13.3 |
| Iron Deficiency Anaemia (<27PG) | 182 | 86.7 |

*Source: Source: Field Survey, 2022*

**Table 9:**
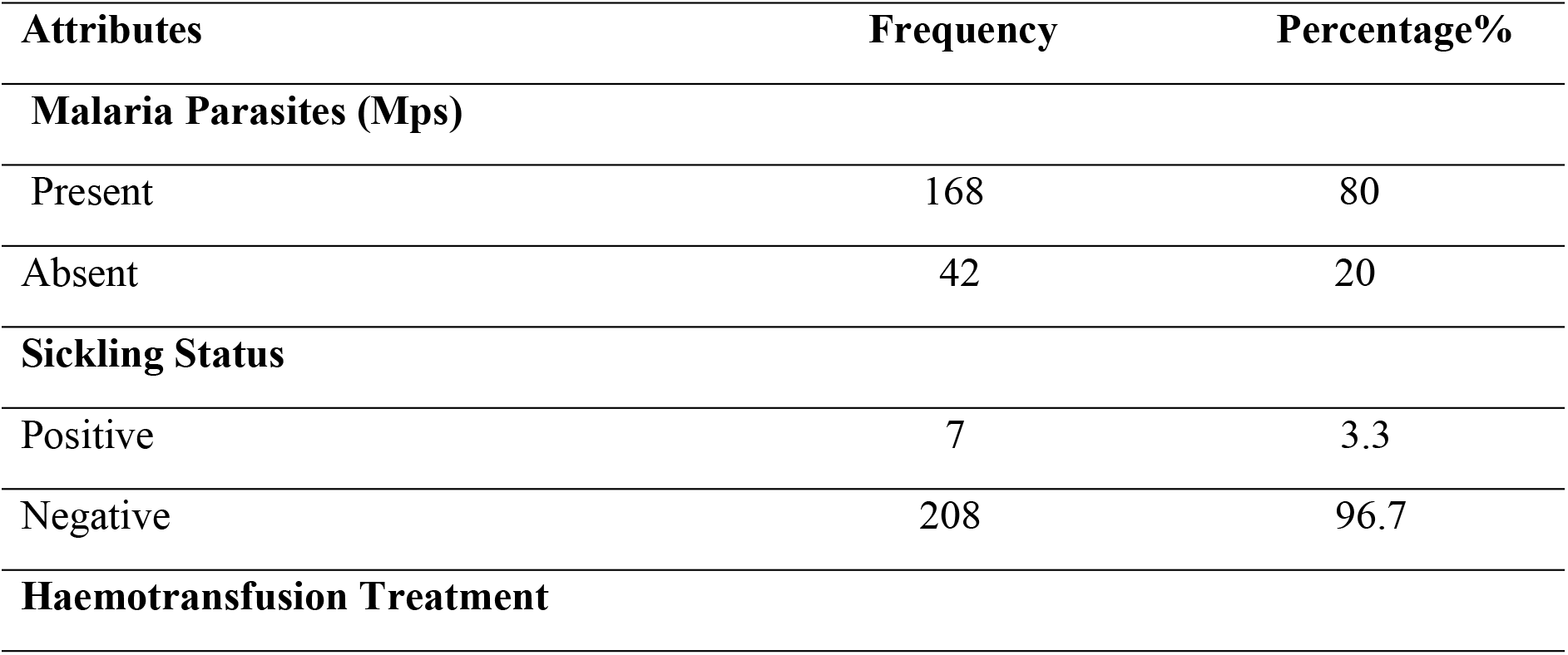
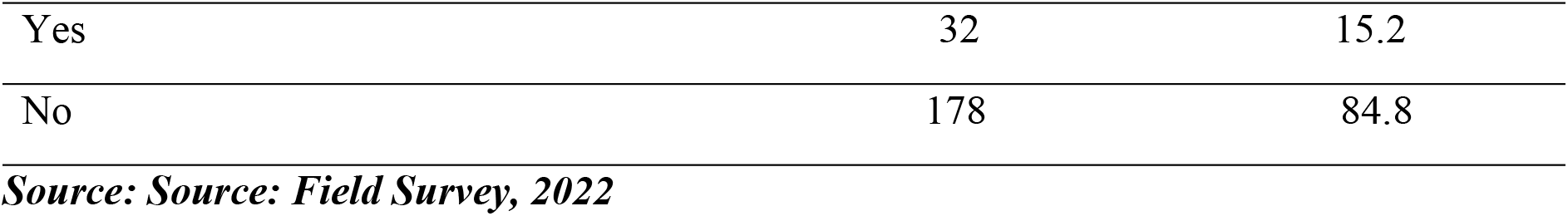
Laboratory investigation (Mps & Sickling) (N=210)

| Attributes | Frequency | Percentage% |
| --- | --- | --- |
| **Malaria Parasites (Mps)** |  |  |
| Present | 168 | 80 |
| Absent | 42 | 20 |
| **Sickling Status** |  |  |
| Positive | 7 | 3.3 |
| Negative | 208 | 96.7 |
| **Haemotransfusion Treatment** |  |  |
| Yes | 32 | 15.2 |
| No | 178 | 84.8 |

***Source: Source: Field Survey, 2022***

The study further observed that 99% of the children had been breastfed while a minority of 1% had not been breastfed. Amongst those who were breastfed, a good majority of 78.1% had been partially breastfed while their counterparts (21%) were exclusively breastfed.

The study additionally sought to find out if any of the children sampled for the survey had any known chronic disease, especially SCD (Sickle Cell Disease) since it is a prerequisite for anaemia, and the study observed that only 3.3% had SCD while a huge majority 96.3% had no known SCD.

Moreover, the study sought to find out the number of times the sampled children were admitted to the hospital in the past one year. The study thus observed that more than half of the children 62.9% of the children had only been admitted once, 21.4% had been admitted twice, 8.6% had been admitted 3 times, 3.8% were admitted 5 times while the least number of the children were admitted 4 times over the previous one year.

The study additionally found that only 41.4% of the children had been dewormed over the past one year while more than half (58.6%) of the children had not been dewormed.

It is a known medical fact that the most common cause of anemia is Iron-Deficiency Anaemia, also lack of vitamin B12 and folate also contribute immensely to anaemia incidence. And since the most consistent source of iron, vitamin B12 and folate is from our diet. The study however sought to probe further, using iron-rich, vitamin B12 rich and folate rich food sources as the basis to find out from guardians how often their children consumed these classes of foods, since they provide the requisite food nutrients which boost the hemoglobin levels and its attendant Erythrocyte levels thereby curbing the menace of anemia. Hence, the lack of these food classes acted as the risk factors to anaemia for the purpose of this current study.

On the consumption of animal protein, the study revealed that majority (39%) of children consumed animal protein more than once in a week, 33.8% consumed it on daily basis, 10.5% consumed it once in a week, 10% indicated not consuming it often while the least 6.7% did not consume it at all.

On the consumption of beans, a majority of 41.4% of children did not consume beans often, 23.8% consumed it more than once in a week, 14.8% of children did not consume this food at all, 13.3% consumed this food once in a week and the least (6.7%) consumed this food on daily basis.

On the consumption of milk, a majority of 30.5% did not consume milk often, 26.2% consumed it more than once in a week, 16.7% did not consume this food at all, 16.2% consumed it on daily basis and the least 10.5% consumed it once in a week.

On the consumption of vegetables, a majority of 32.4% of children consumed vegetables more than once in a week, 27.1% did not consume it often, 15.2% consumed it once in a week, 14.3% of children did not consume vegetables at all while 11% of children consumed vegetables on daily basis,

On the consumption of fruits, a majority of 37.1% of children consumed fruits more than once in a week, 19% consumed it once in a week, 18.1% did not consume it often, 15.7% of children consumed fruits on daily basis while 10% did not consume fruits at all.

On the consumption of composite porridge, a majority of 36.7% of children did not consume composite porridge often, 29.5% consumed it more than once in a week, 15.2% consumed it once in a week, 10% of children consumed composite porridge on daily basis while 8.6% of children did not consume composite porridge at all.

Lastly, on the consumption of tea with sugar, the study observed that 35.7% of children consumed tea with sugar more than once in a week, 25.2% did not consume it at all, 17.6% did not consume if often, 13.3% of children consumed tea with sugar on daily basis while 8.1% consumed tea with sugar once in a week.

In order to effectively draw scientific and medically certified conclusions about the anaemia situation in Sekyere East district and its environs, principal diagnosis conducted on the sampled children by qualified medical practitioners were collated together to ascertain the status of the children. However, for the purpose of this Anaemia-oriented survey, strict emphasis was placed on anemic related conditions.

The study observed that 15 children (7.1%) were diagnosed of severe anaemia, 6 children (2.9%) were diagnosed of moderate anaemia, 3 children (1.4%) were diagnosed of mild anaemia.

The study further observed that, 1 child (0.5%) was diagnosed of mild anaemia and sepsis concurrently, another 1 child (0.5%) was diagnosed of severe anaemia and severe malaria concurrently. Also, another 1 child (0.5%) was diagnosed of moderate anemia and sepsis concurrently, 2 children (1%) were diagnosed of moderate anaemia and malaria concurrently, another 2 children (1%) were diagnosed of Moderate anaemia and Severe malaria concurrently.

Also, 5 children (2.4%) were diagnosed of severe anaemia in Sickle Cell Disease (SCD) and another 3 children (1.4%) were diagnosed of Mild anaemia and Severe malaria concurrently.

Findings from the principal diagnosis as presented above revealed that 39 of the children representing 18.7% were diagnosed of anaemia and other anemic related associated conditions.

The study further took into consideration all cases of malaria, and this is due to the fact that malaria can be a precursor to developing anaemia since during the infectious stage of its life cycle, these parasites invade the red blood cells in an attempt to evade the human immune system. After the invasion, the malaria parasites remodel the red blood cells for their own use thereby leading to anemia situations.

The study therefore observed that, 63 of the sampled children (30%) were diagnosed of severe malaria, 42 children (20%) were diagnosed of malaria, 5 children (2.4%) were diagnosed of severe malaria and febrile convulsion concurrently, 1 child (0.5%) was diagnosed of severe malaria and UTI concurrently, 4 children (1.9%) were diagnosed of malaria and Gastroenteritis concurrently.

Also, 2 children (1%) were diagnosed of malaria and sepsis concurrently, another 2 children (1%) were diagnosed of severe malaria and enteritis and 1 child (0.5%) was diagnosed of malaria in Sickle Cell Disease (SCD).

After a thorough blood analysis of the sampled 210 children enrolled for the survey, the haemoglobin levels vividly painted a clear picture of the situation of anaemia in Sekyere East. The study observed that little above half (124) children representing 59.0% had normal hemoglobin levels which is indicative of the absence of anaemia.

However, 72 of the children (34.3%) had haemoglobin levels indicative of mild/moderate anaemia while 14 of the children (6.7%) had haemoglobin levels indicative of severe anaemia.

This study therefore revealed that 86 of the sampled children representing 41% of the children had confirmed varying forms of anaemia.

In order to spice up the Anaemic situation revealed by the Haemoglobin (Hb) tests, Mean Corpuscular Volume (MCV) test was done to measure the average size of the red blood cells of the sampled children.

The study however observed that only 33 (15.7%) had normal Mean Corpuscular Volume levels while a huge majority of the children 176 (83.8%) had abnormal Mean Corpuscular Volume levels which was suggestive of iron-deficiency anemia. Interestingly, one child had a Mean Corpuscular Volume level above 95FL which was suggestive of Vitamin B12 or folate deficiency anaemia.

In order to place the anaemia situation in Sekyere East in its right perspective, a Mean Corpuscular Haemoglobin blood analysis were run on blood samples taken from the sampled children. MCH levels basically refer to the average amount of haemoglobin in the red blood cells in the body.

The study observed that only 28 (13.3%) of the children had normal MCH levels while a greater majority 182 (86.7%) had abnormal levels of MCH suggestive of Iron-Deficiency anaemia.

Based on the enormous danger malaria parasites pose to red blood cells during their life cycle and its significant contribution to most anaemia cases, the researchers thought it wise to run a laboratory-based blood analysis on the blood samples of the children enrolled for the survey.

The study observed that a greater majority of the children 168 (80%) tested positive for the malaria parasites and this may have accounted for the huge anaemia numbers in the Hb, MCV and MCH blood analysis findings.

The study additionally sought to find out the sickling status of the children enrolled for the study since Sickle Cell Disease and Anaemia move together. The study thus observed that, only 7 (3.3%) of the children had Sickle Cell Disease while a greater proportion of the children 203 (96.7%) were negative for the Sickle Cell Disease.

Haemotransfusion is a treatment measure to shore up the haemoglobin levels of patients who are anemic. Hence, the principal researchers sought to find out how many of the sampled children got blood transfusion as a medical response to remedy the anemic condition. The study found that 32 (15.2%) of the children were haemotransfused while their majority counterparts of 178 (84.8%) were not.

## 4.0 DISCUSSION

### 4.1.1 Prevalence of anaemia in Sekyere East district

The Haemoglobin(Hb) blood analysis of the study revealed that 86 of the sampled children representing 41% of the total children had confirmed varying forms of anaemia (mild/moderate and severe) in the Sekyere East district.

Moreover, the MCV blood analysis revealed that a huge majority of the children 176 (83.8%) had abnormal Mean Corpuscular Volume levels which was suggestive of iron-deficiency anemia. Interestingly, one child (0.5%) had a Mean Corpuscular Volume level above 95FL which was suggestive of Vitamin B12 or folate deficiency anaemia. Therefore, the MCV analysis revealed 177 (84.3%) cases of anaemia in the Sekyere East district.

The Mean Corpuscular Haemoglobin (MCH) test further revealed that a greater majority 182 (86.7%) of the sampled children had abnormal levels of MCH suggestive of Iron-Deficiency anaemia.

The above findings reflect the anaemia situation in the Sekyere East district, this however calls for concerted efforts between major stakeholders to arrest the childhood anaemia canker that is gradually gaining grounds in the district. From the public health perspective, this situation needs every strategic arsenal to mitigate this childhood anaemia menace at its benign stage before it attains malignancy since it can be a major trigger of morbidity and mortality.

These findings however corroborate and affirm the observations made by Ewusie, Ahiadeke and Beyene (2022) who opined that the prevalence of anaemia among Ghanaian children is high (74.8%) in their study where they did a systematic review of the Ghana Demographic and Health Survey.

This also study also confirm similar findings by Egbi et al (2021) who assessed the prevalence of anaemia among school going pupils in Volta region and reported high prevalence of anaemia among the children.

These findings are also in tangent with similar observations made by Nambiema, Alexie & Issifou (2019) who observed high prevalence of anaemia (70.9%) among children in Togo after their systematic review of Togo demographic and health Survey data 2013-2014.

### 4.1.2 Malaria and Anaemia

The study observed that a greater majority of the children 168 (80%) tested positive for the malaria parasites and this may have accounted for the huge anaemia numbers in the Hb, MCV and MCH blood analysis findings.

According to Hung et al, 2022, anaemia has been shown to be prevalent in areas where malaria is endemic. This study however corroborates the findings from Hung et al (2022) due to the high positive malaria cases among the children enrolled for the study. Malaria parasites are involved in increased destruction and reduced production of red blood cells thereby inducing anaemia.

### 4.1.3 Risk factors for Anaemia in Sekyere East district

It is a known medical fact that the most common cause of anemia is Iron-Deficiency Anaemia, also lack of vitamin B12 and folate also contribute immensely to anaemia incidence. And since the most consistent source of iron, vitamin B12 and folate is from our diet. The study however sought to probe further, using iron-rich, vitamin B12 rich and folate rich food sources as the basis to find out from guardians how often their children consumed these classes of foods, since they provide the requisite food nutrients which boost the hemoglobin levels and its attendant Erythrocyte levels thereby curbing the menace of anemia.

Premising on the aforementioned paragraph, animal protein, beans, milk, vegetables, fruits, composite porridge and tea with sugar (which are very rich in iron, vitamin B12 and folate) were selected as the indicators for good dietary intake among the sampled children.

However, on the basis of comparative analysis grounded on percentages, the study observed moderate consumption rate (not too high and not too low) of these nutrient rich food products necessary to curb anaemia.

According to Nevins (2008), poor child nutrition leads to anaemia and this study affirms same and this is due to the fact that, the moderate to average intake of these vital food products may have accounted for the high prevalence of anaemia among the sampled children.

### 4.1.4 Frequency of Haemotransfusion among children in Sekyere East

The study found that 32 (15.2%) of the sampled children were haemotransfused due to their anemic conditions.

Red blood cell (RBC) transfusions are a mainstay in the treatment of anemic patients, making it the most common medical procedure in hospitalized patients. Most RBC transfusions are prescribed for patients with relatively low levels of hemoglobin (Hb) and only in controlled situations. The underlying thinking is that the transfusion will increase oxygen transport and therefore decrease deficiencies thus relieving tissue hypoxia (inadequate oxygen).

According Buelvas (2021), an inadequate supply of oxygen to tissues can lead to multiple organ failure and increased morbidity and mortality. These deleterious effects appear only with very low Hb levels when compensatory mechanisms do not work properly or are insufficient and this current study reasons in the same direction.

## 5.0 CONCLUSIONS

### 5.1 Conclusions

This current study has accentuated high prevalence of anaemia in the Sekyere East district looking at the various findings brought to light by the Haemoglobin (Hb) blood analysis, Mean Corpuscular Volume (MCV) blood tests and the Mean Corpuscular Haemoglobin (MCH) test results.

The findings from the study have also shown that malaria in children is significantly associated with the onset of childhood anaemia.

The study additionally revealed moderate nutritional intake of food products very rich in iron, vitamin B12 and folate nutrients necessary to curb anaemia in children.

In a nutshell, study findings therefore underscore the need for multi-pronged approaches that address both malaria control and nutrition in order to reduce anaemia among the children in Sekyere East district.

### 5.2 Recommendations

Based on the findings and conclusions from the study, the following recommendations are suggested

1. Maternal education should be highly encouraged. A higher level of maternal education leads to increased knowledge about health and nutrition, which, in turn, leads to an increase in the quality of the diets consumed by children. The district health directorate, the municipal assembly and the district hospital should spearhead this vital campaign targeted at dealing with anaemia.
2. There should be a district policy tailored along anaemia lines, to give iron fortified supplements to children in the Sekyere East district. This when effectively and adequately done would contribute immensely towards the drastic reduction in childhood anemia.
3. The malaria control programme in the Sekyere East district should be highly strengthened by the Municipal Assembly and the District Health Directorate. This can be achieved through the addition of logistical support where necessary to catalyze the objective of the programme i.e. bringing malaria prevalence down. This when effectively done would lead to the distribution of insecticide treated nets and mass malaria awareness campaigns which will all culminate in the drastic reduction in childhood malaria. This would invariably lead to the reduction in childhood anaemia cases in the Sekyere East District.

## Data Availability

Data will be made readily available upon request by the journal

## Acknowledgement

Our greatest heartfelt appreciation goes to dad Prof. Kofi Bobi Barimah, Mrs. Sandra Adelaide Hanson and Nana Ama Frimpomaa for their spectacular help in diverse ways. We also acknowledge the Director of the College of Health Yamfo Dr. Dr Mohammed Mohammed Ibrahim for his visionary leadership in the area of research.

